# An environmental determinant of viral respiratory disease

**DOI:** 10.1101/2020.06.05.20123349

**Authors:** Yeon-Woo Choi, Alexandre Tuel, Elfatih A. B. Eltahir

## Abstract

The evident seasonality of influenza suggests a significant role for weather and climate as one of several determinants of viral respiratory disease (VRD), including social determinants which play a major role in shaping these phenomena. Based on the current mechanistic understanding of how VRDs are transmitted by small droplets, we identify an environmental variable, Air Drying Capacity (ADC), as an atmospheric state-variable with significant and direct relevance to the transmission of VRD. ADC dictates the evolution and fate of droplets under given temperature and humidity conditions. The definition of this variable is rooted in the Maxwell theory of droplet evolution via coupled heat and mass transfer between droplets and the surrounding environment. We present the climatology of ADC, and compare its observed distribution in space and time to the observed prevalence of influenza and COVID-19 from extensive global data sets. Globally, large ADC values appear to significantly constrain the observed transmission and spread of VRD, consistent with the significant coherency of the observed seasonal cycles of ADC and influenza. Our results introduce a new environmental determinant, rooted in the mechanism of VRD transmission, with potential implications for explaining seasonality of influenza, and for describing how environmental conditions may impact to some degree the evolution of similar VRDs, such as COVID-19.

---

The spread of Viral Respiratory Diseases (VRDs), like influenza and COVID-19, is shaped by a combination of social, biological and environmental determinants. Social determinants include behavioural aspects (settlement density, mobility, personal hygiene, vaccination, social distancing, etc.) that affect the transmission of the disease. Biological determinants are defined here as characteristics of the pathogen itself including its response to abiotic factors, and the nature of the human immune response to it. Finally, environmental determinants are defined here as the set of environmental conditions that impact the intensity of the disease transmission process. (For example, how much a virus tolerates extreme temperature is a biological determinant, while how extreme temperature impacts the transmission of the virus is an environmental determinant.) Most public policy approaches to limit the spread of VRDs typically rely on manipulating social behaviours through emphasis on personal hygiene, social distancing and vaccination, and COVID-19 is no exception. Still, the dynamics and average prevalence of VRDs exhibit substantial variability across countries. Influenza is most widespread in the mid-latitudes,^1^ and in the case of COVID-19, some countries have clearly experienced widespread transmission and an explosive growth in cases, while in others, the outbreak seems much more constrained.^2–4^ It is evident that social determinants play a major role in controlling transmission, especially given the success of social distancing policies implemented in response to COVID-19. However, this does not necessarily imply that the environment plays no role in shaping VRD spread, as highlighted by the clear seasonality of influenza in mid-latitude countries.^1^

We still do not have a definite understanding of the biological determinants of VRDs. Laboratory experiments have suggested that ambient temperature and absolute humidity affected the survival of several VRD pathogens,^5–7^ although the effect of temperature seems weak in the case of the SARS-CoV-2 virus responsible for COVID-19^8^ (Fig. S1). High UV radiation is also believed to suppress viral activity and infectivity in the case of influenza viruses^9^ and possibly SARS-CoV-2.^10^ Additionally, evidence has emerged that viral shedding in mammals is enhanced at low temperatures,^11^ making the case for strong biological controls on VRD prevalence. Yet, because VRDs are primarily transmitted by droplets exhaled by infected subjects, environmental conditions may also play a major role in shaping their spread.^12,13^ Previous studies have argued that cold and dry environments were conducive to the survival and transport of VRD-infected droplets, unlike warm and humid environments.^1,6^ This hypothesis seems supported by empirical relationships applied to country-level data,^7,14,15,16^ though in the case of COVID-19 initial results suggest that weather and climate conditions may have limited effects on the spread of the disease.^17,18^

One important limitation of such studies is their focus on temperature or humidity as separate covariates to understand or predict VRD prevalence. Different relationships are developed for tropical and mid-latitude countries^1^ although the physics of droplets is the same. Additionally, relationships are sometimes found to be non-monotonic: in the case of COVID-19, the transmission efficiency may first be enhanced as temperature and absolute humidity drop, and then decline beyond a certain threshold.^15^ Therefore, while evidence points to some degree of environmental control on VRD spread and prevalence, the lack of a consistent and physically-based framework makes it all the more difficult to assess.

Here, we propose a new atmospheric state-variable, named Air Drying Capacity (ADC), rooted in the current mechanistic understanding of how transmission takes place by small droplets. ADC is defined as the rate of decrease with time of a droplet surface area, given ambient temperature and humidity. As such, ADC integrates naturally the effects of both temperature and humidity based on their relative roles in dictating the decrease of the droplet surface area. ADC offers a consistent, physically-based framework to assess the effect of environmental conditions on global and seasonal patterns of VRD prevalence.

## Methods

### Droplet Theory of VRD Transmission

VRDs are believed to be transmitted by droplets exhaled by infected subjects.^19–22^ The size of exhaled droplets by human typically ranges from about 0.5 µm in breathing, and increases with speech up to about 10 µm.^23^ Larger droplets, up to 200–300 µm, can be emitted by sneezing or coughing.^24^ After emission, droplets can contaminate nearby surfaces, or disperse as aerosols and may infect subjects who inhale them. This idea, first developed by Wells,^25^ led to the discrimination of “large” and “small” droplets, and has since then influenced strategies to control the spread of infection according to whether the disease was thought to be transmitted primarily through large or small droplets.^26^ Droplet diameter cut-offs usually range between 5 and 10µm,^27^ and the typical associated distance varies between 1.5 and 2m.^28^ More recent studies have shown, however, that these arbitrary droplet size cut-offs do not reflect the actual trajectories of exhaled droplets. The dynamics of droplet evaporation and evolution are indeed very dependent on the characteristics of the complex multiphase turbulent flow which the droplets exist in^29^ as well as background environmental conditions.^12^ While influenza transmission has been shown to occur through both the large and small droplet route,^21^ at this stage, COVID-19 is still believed to be mainly transmitted by the large droplet path,^30^ although aerosol transmission may also be possible.^31^ In any case, exhaled droplets are still the major infection route, which implies that environmental controls on droplet evaporation and disappearance may play an important role in determining the spread of the disease.

### A proposed environmental determinant: Atmospheric Drying Capacity (ADC)

Droplet growth theory under given environmental conditions goes back to the pioneering work of Maxwell,^32^ who first posited that steady-state dynamics of spherical droplets at rest in isotropic gaseous media were controlled by the equilibrium between heat and mass exchange at their surface. Both mass and heat transfer involve ambient temperature and humidity, and are therefore strongly constrained by environmental conditions. In steady-state, mass and heat transfer exactly compensate, and one finds that the radius *r* of a droplet evolves according to^33^:

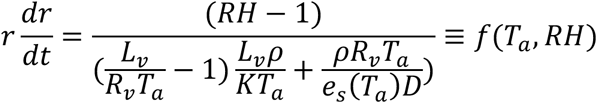

where *RH* is the ambient relative humidity, *T_a_* the ambient temperature, *R_v_* the specific gas constant for water vapour, ρ liquid water density, *L_v_* the latent heat of vaporisation, *K* the thermal conductivity of air, *D* the water vapour diffusion coefficient, and *e_s_(T)* the saturation vapor pressure at temperature *T* given by the Clausius-Clapeyron equation. We then define the Air Drying Capacity (ADC, in mm^2^/hr) as the rate of decrease of the droplet surface area:

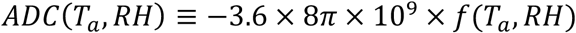

ADC is therefore an atmospheric state-variable uniquely related to air temperature and humidity only. For typical ranges of air temperature and humidity, ADC varies between 0 and 15 mm^2^/hr (Fig. 1-a,b). It is a linear function of both relative and specific humidity, but a non-linear function of temperature, consistent with the Clausius-Clapeyron law. ADC strongly controls the time it takes for a free-falling droplet to evaporate, and therefore the diameter cut-off between “large” droplets, that reach the ground before evaporating, and “small” droplets, which turn into aerosols (Fig. 1-c). At low ADC values (0–1 mm^2^/hr), only droplets larger than about 25µm will be able to contaminate nearby surfaces, while for high ADC (>10 mm^2^/hr) that threshold moves up to 60µm. Additionally, the potential range of such large droplets is also severely reduced as ADC increases, because they can remain in the air for a significantly shorter time (Fig. 1-c, Fig. S2). Small (<10µm) droplets - a size typically emitted during normal speech - while never able to contaminate surfaces under the typical range of ADC values, can however potentially be inhaled by subjects in the vicinity of the emitter. Their fate is largely controlled by ADC: a 10µm droplet will evaporate in as much as 25s or as less as 0.5s depending on the background ADC. This may be particularly relevant for VRD pathogens whose infectivity declines once in the dry aerosol phase.

**Figure 1.**
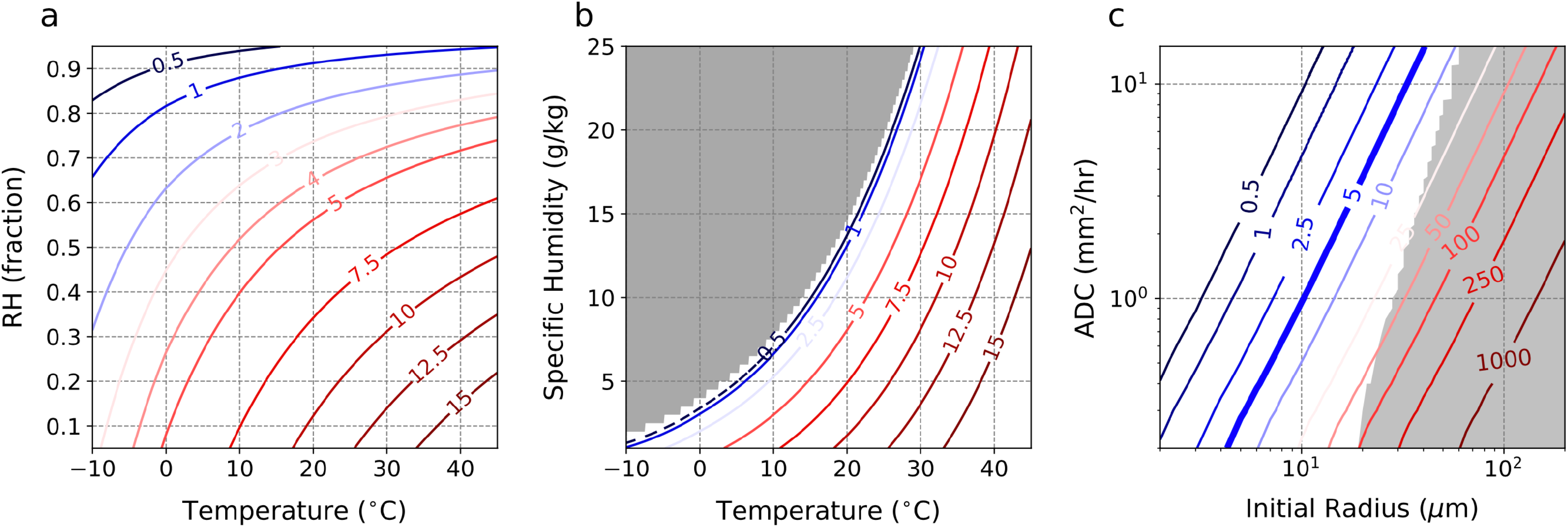
ADC and environmental conditions. (a-b) Air Drying Capacity (ADC, unit: mm^2^/hour) as a function of temperature and (a) relative humidity and (b) specific humidity. The grey area in (b) indicates super-saturation. (c) Time to evaporation of free-falling, spherical water droplets as a function of ADC and initial droplet radius. The shaded area indicate the region where droplets reach the ground before evaporating.

### Data

6-hourly temperature, dew point temperature and surface pressure data at 0.75° spatial resolution between 1979 and 2018 were obtained from the ERA-Interim reanalysis^34^ (available at http://apps.ecmwf.int/datasets/). Since ERA-Interim is only available up to 2019, we used 6- hourly ERA5T^35^ data (0.25° × 0.25° horizontal resolution) for the recent February-April 2020 period.

Daily COVID-19 epidemiological data compiled by the Johns Hopkins University Center for Systems Science and Engineering is available at country-scale since 22 January 2020 at https://data.humdata.org/. The most up-to-date COVID data for each US state at a daily temporal resolution is taken from the COVID Tracking Project (https://covidtracking.com/data/). Population data for world countries and US states was downloaded from https://www.worldometers.info/world-population and https://www.wikipedia.org, respectively. Weekly laboratory confirmed influenza cases by country for the period October 15^th^, 1995 to August 31^st^, 2019 are retrieved from the World Health Organization’s FluNet database, accessible at https://www.who.int/influenza/gisrs_laboratory/flunet/en/. The dataset consists in weekly totals of identified influenza A and B cases, along with the number of subjects tested. It suffers from both a sampling bias (variations with time and space in the number of people tested), and a reporting bias (reports are not available consistently over time). We use two indices of influenza prevalence to separately address these biases. First, we define an “influenza frequency index”, defined weekly as the number of positive influenza A and B cases divided by the number of tested subjects. Second, we define a “normalized influenza prevalence (NIP)” index based on the approach of Deyle et al.^14^ as the number of positive cases divided by population (linearly interpolated over time to account for population trends), and multiplied by the average number of annual reports for all countries divided by the average number of annual reports for the country in question:

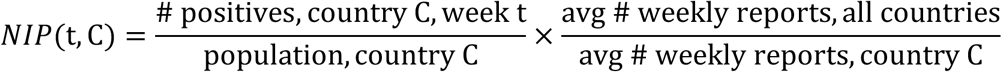

(see supplementary methods for more details).

## Results

### Climatology of ADC

The spatial distribution of annual-average ADC shows a somewhat meridionally symmetric pattern. The lowest values, between 0–2 mm^2^/hr, can be found above 60° latitude in each hemisphere and over land areas around the equator (Fig. 2-a). The subtropics in each hemisphere exhibit high ADC values, particularly over the large deserts of North Africa and southwest Asia where temperature is high and humidity is low. Australia, India and the Western United States are all characterised by relatively high ADCs. The situation during winter and spring is overall quite similar, though with notable regional differences (Fig. 2-b,e). Europe and Eastern North America both show particularly low ADC values during winter, much lower than in China where ADC remains mostly above 2 mm^2^/hr. ADC over south-eastern Brazil is also at its minimum (Fig. 2-e). By contrast, most of Africa, and specifically its large population centres of Ethiopia, Egypt and Nigeria, all show high ADCs. The same can be said for India, particularly during spring. However, consistent with the summer monsoon cycle, ADC becomes much higher during and after the monsoon season over Western Africa and the Sahel region, as well as India, as high-ADC bands move northwards with the rains (Fig. 2-c,d). Over Western Europe and Eastern North America, ADC increases during summer, but remains rather low at around 5 mm^2^/hr. A video showing the space-time evolution of ADC is included with Supplementary Information.

**Figure 2.**
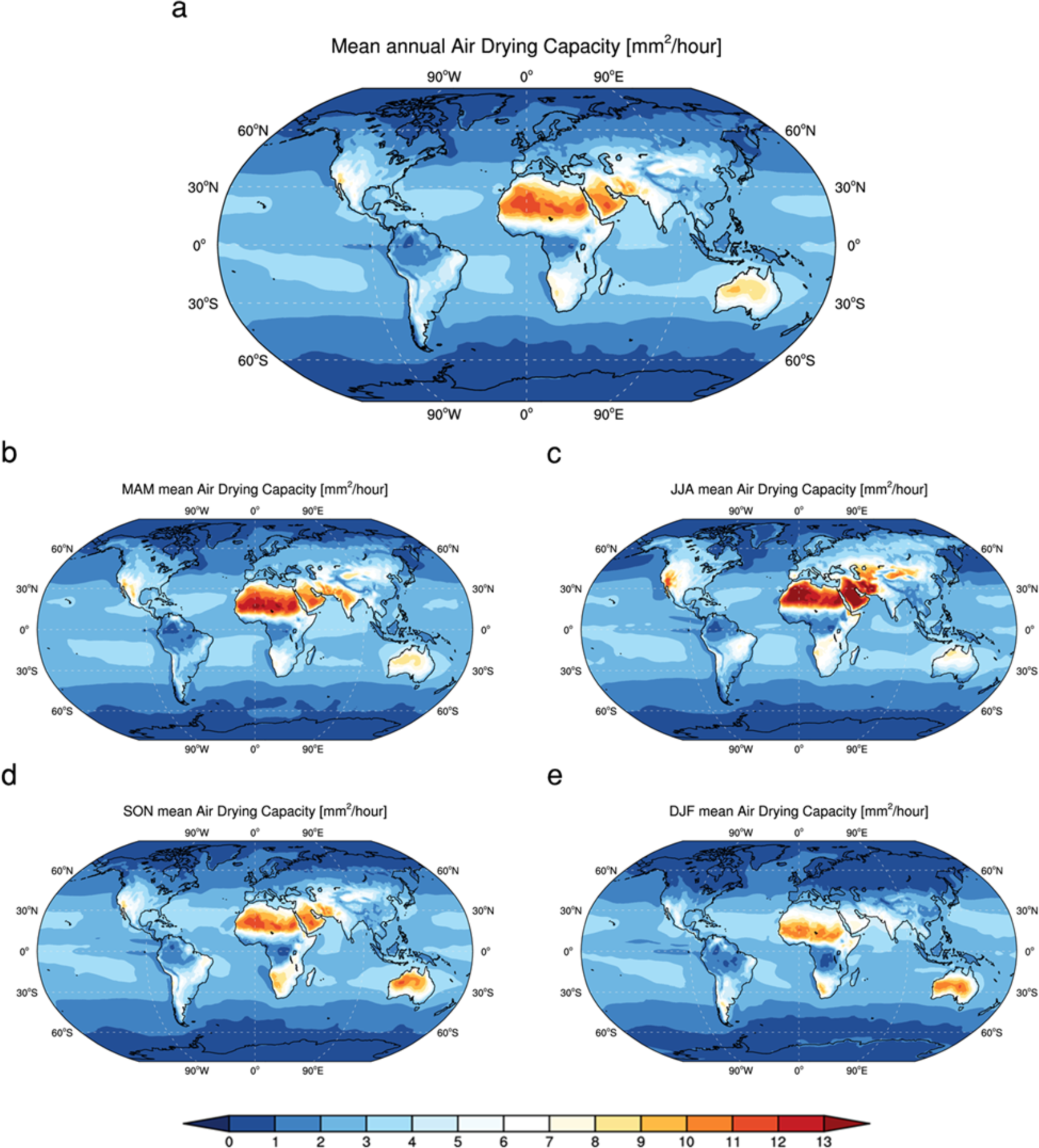
Global distribution of ADC. (a-e) Global map of (a) annual, (b) spring (March to May), (c) summer (June to August), (d) autumn (September to November), and (e) winter (December to February) average ADC for the period 1979–2018, calculated from the ERA-Interim dataset.

### Testing the relevance of ADC for VRD prevalence

The spatial and temporal distribution of influenza cases is highly consistent with that of ADC (Figs. 3-a, 4-a,b). ADC appears to set a strong upper bound on influenza prevalence that applies to all countries with available data: influenza has very limited prevalence at ADCs of 5 mm^2^/hr or larger, and clearly increases as ADC approaches 0 (Figs. 3-a, 4-a,b). The annual cycles of ADC and influenza are also highly consistent, with a clear peak in the disease around when ADC is at its lowest (Fig. 3-c). Africa stands out due to high ADC values and low influenza prevalence, whereas Europe and North America have low ADCs and generally higher numbers of influenza cases (Fig. 4-a,b). While socio-economic factors also play a role in modulating the spread of the disease, it is striking that ADC still constrains the upper end of the range of observed prevalence, consistent with its effect on droplets – the vectors of transmission, particularly the rapid increase in the time needed for droplet evaporation as ADC approaches 0 (Fig. S2). By contrast, air temperature (Fig. 3-b) and specific humidity (Fig. S3-a,c) do not show such clear relationships to influenza, although the annual cycle of temperature appears quite consistent with that of influenza prevalence (Fig. 3-d). Results for relative humidity do show some enhancement of influenza as the air becomes moister (Fig. S3-b), but its annual cycle seems quite off when compared to that of influenza incidence (Fig. S3-d).

**Figure 3.**
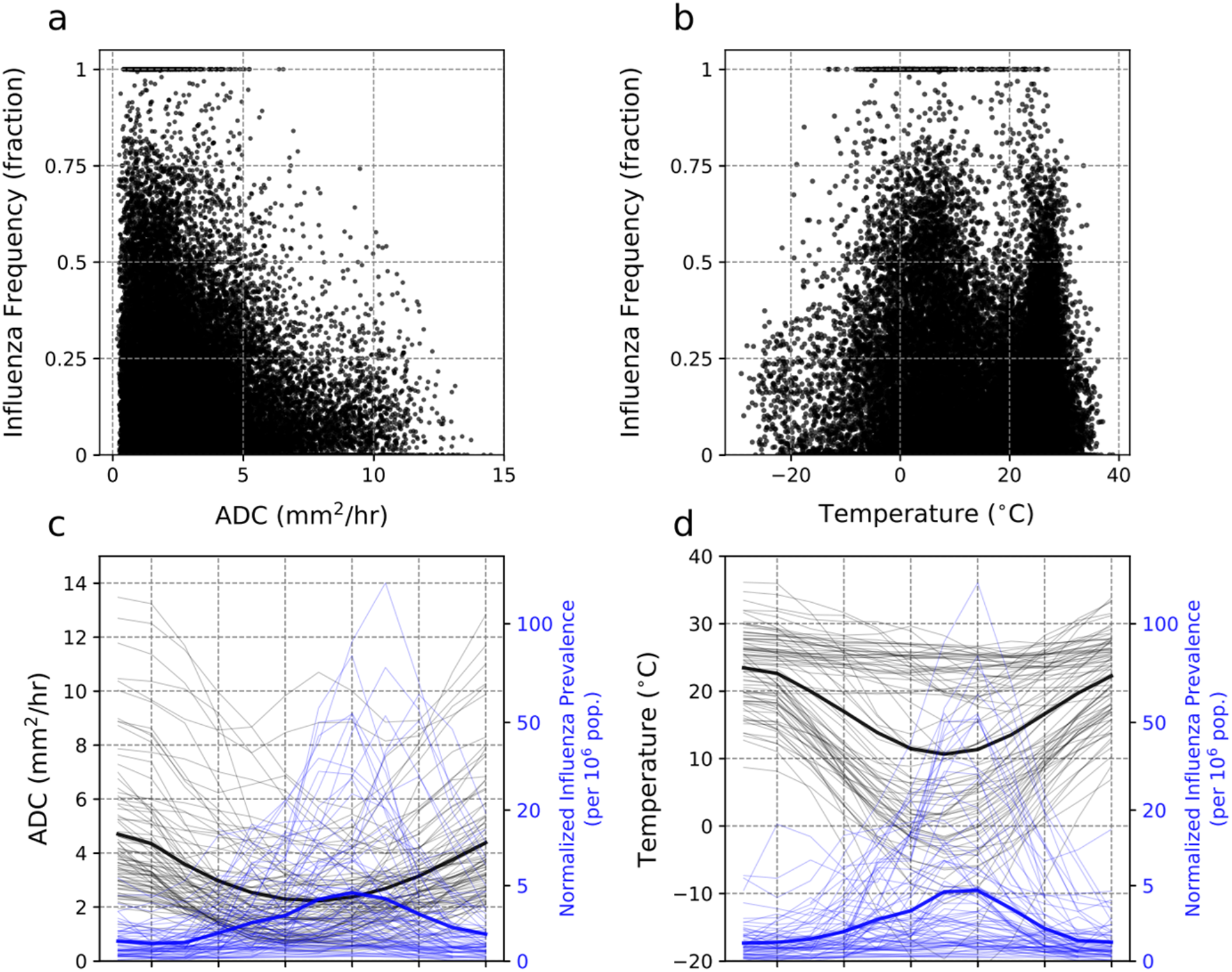
Seasonal variation of ADC and prevalence of VRD. (a-b) Weekly influenza frequency against (a) ADC and (b) temperature. (c-d) Seasonal variation of normalised influenza prevalence alongside (c) ADC and (d) temperature. In (c-d), the first month is defined for each of the 85 countries as the month with maximum ADC (c) or temperature (d).

**Figure 4.**
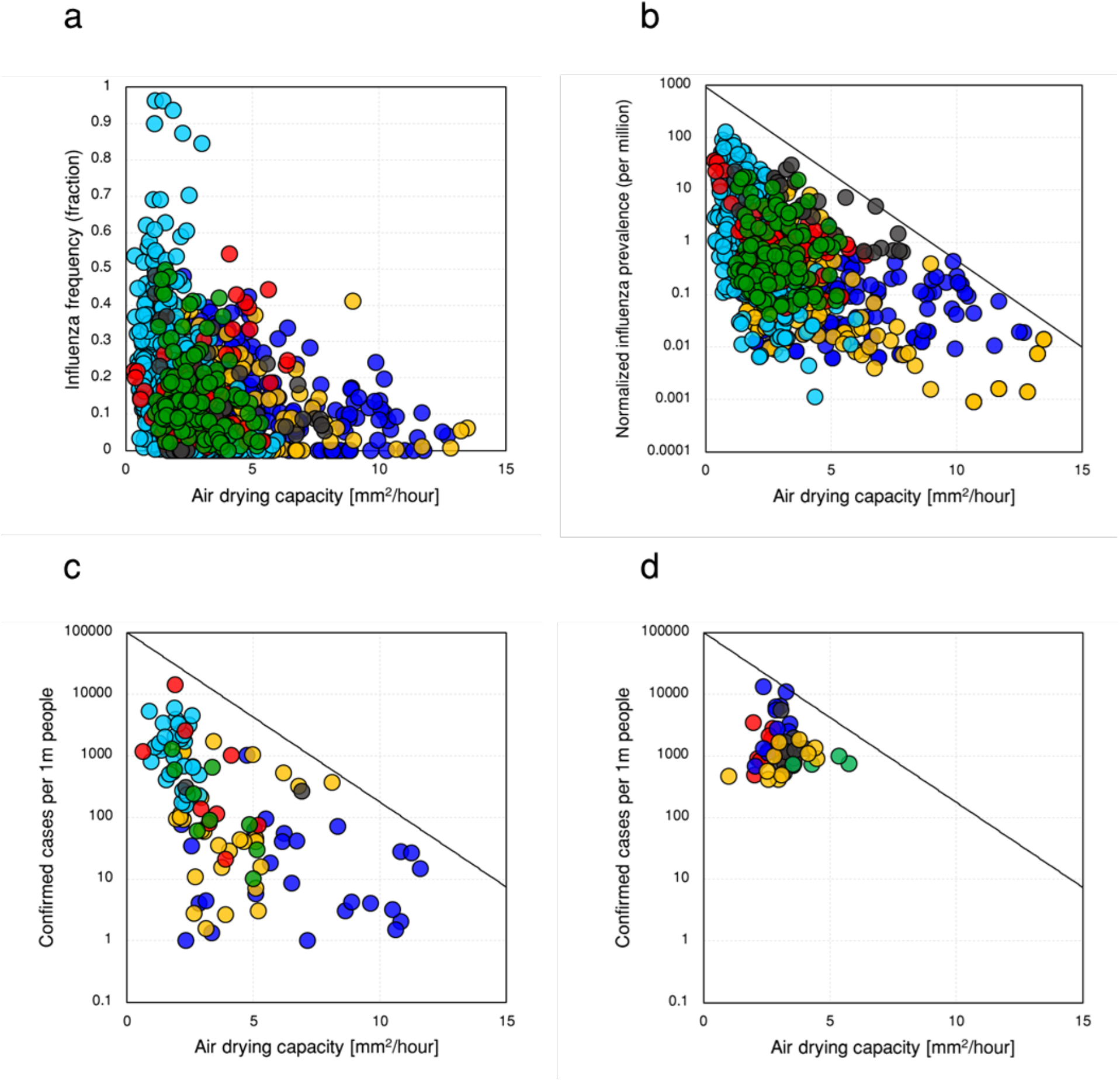
ADC and viral respiratory disease (VRD) prevalence. (a-b) Long-term monthly-mean ADC against long-term monthly-mean (a) influenza A and B frequency, and (b) normalized influenza pervalence for 85 countries (Table S1) for the 1995–2019 period. (c) February-April 2020 ADC against concurrent accumulated confirmed COVID-19 cases for 108 countries (Table S1). (d) Same as (c), but for the 50 US states. Red, green, light blue, yellow, blue and black colors in (a,b,c) respectively indicate North America, South America, Europe, Asia, Africa, and Oceania countries, and in (d) the Western, North-eastern, Midwestern, Southeastern, and South-western US states are represented by yellow, blue, red, black, and green colours, respectively.

Interestingly, the spatial distribution of ADC during winter and spring also shows some resemblance to the global map of confirmed COVID-19 cases (Fig. 4-c,d, Fig. S4). The disease hotspots of Europe and Eastern North America (>1000 cases per million) both have extremely low ADCs, whereas China and the Western United States have fewer cases per million and larger ADC, despite also being highly connected to the rest of the world. COVID-19 prevalence in South America and Australia is lower (10–500 per million), and even less than that in Africa and India. Naturally, many other factors come into play here, like connectivity to the rest of the world, population density and localisation within countries, and public policy measures like social distancing or lockdowns. The number of reported cases also suffers from biases, especially undercounting. Still, countries with low (respectively high) ADCs generally seem to correspond to higher (respectively lower) disease prevalence, a tendency that seems robust to considerations of income levels or test numbers performed by different countries (Fig. S5).

## Discussion and Conclusions

VRDs are primarily transmitted between humans through droplets exhaled by infected hosts. Environmental determinants that affect the fate of these droplets can therefore influence transmission of these diseases. We introduced here a new variable, ADC, motivated by droplet growth theory first developed by Maxwell^32^. ADC includes the effects of both temperature and humidity on droplet evolution in the atmosphere. Compared to temperature, ADC turns out to set a much more coherent constraint on influenza prevalence (Fig. 3). The empirical relationship of ADC with the average prevalence of both influenza and COVID-19 for various world regions is consistent with its physical effects on the decay of droplets through which VRDs are transmitted. ADC directly constrains the evaporation of airborne droplets, potentially setting a strong upper bound on VRD spread and prevalence that appears valid regardless of socio-economic factor (Figs. 4, S5). It is important to note that ADC also indirectly impacts the survival of liquid droplets even once they have landed on surfaces; high-ADC conditions lead to rapid evaporation from a surface. The transmission of the viruses responsible for COVID-19 and influenza is thus likely impacted by ADC.

Significant relationships between temperature or humidity and influenza dynamics have been suggested in previous studies for individual countries^14^ and temperate regions^6^, but it appears that neither variable, unlike ADC, is able to explain the observed global pattern of influenza prevalence (Figs. 3, S3). In particular, while specific humidity seems to have a strong effect on influenza virus survival, potentially affecting its transmission during the relatively low-specific humidity peak season in mid-latitude countries^6^, peak influenza in different countries occurs at both times of minimum and of maximum specific humidity^1^. The environmental determinant of VRD proposed in this study has important implications for consistently explaining the seasonality of influenza across the globe. Two kinds of favourable environments have been suggested for influenza transmission: “cold-dry” (as in mid-latitude countries) and “humid-rainy” (as in tropical countries),^1^ in order to reconcile discrepancies in explaining seasonality of influenza at the global scale.^36^ However, if specific humidity were the determinant variable impacting transmission, humid countries would hardly experience any influenza outbreaks, especially during their wet season. Two clusters of high influenza prevalence can be found in the WHO dataset, at both very low and very high humidity (Fig. S6). What they have in common are low ADC values, and in fact each cluster corresponds to the period of annual minimum ADC in mid-latitude and in tropical countries. The two proposed influenza regimes may therefore be reconciled by considering ADC framework proposed in this paper. While humidity and temperature may mimic influenza dynamics at the scale of individual countries,^6,14^ these same relationships seem less valid when assumed for the world as whole and do not explain the large discrepancies in influenza prevalence between countries. At the global scale, it appears that the environment’s direct effect on droplets, the VRD transmission vectors, and described here using ADC, dominates over its biological effect on virus survival.

While ADC may only set an upper limit to VRD prevalence, social determinants like individual behaviour, socioeconomic conditions, healthcare expenditure, population density, cultural norms, etc. play a major role in shaping such diseases, and likely explain much of the spread in VRD prevalence below the ADC-dependent threshold. Strictly speaking, ADC describes the environmental conditions under which VRDs are likely to be transmitted. Whether or not transmission actually occurs depends, in addition to ADC, on several complex biological as well as social factors. As demonstrated in Figs. 3 and 4, the same value of ADC corresponds to a range of values of observed cases of VRD. That variability in spread is undoubtedly linked to the social and biologic factors independent of ADC, as well as the history of the disease in that location including seeding from other locations. However, the upper limit on the observed range of prevalence decreases over several orders of magnitude as ADC increases, highlighting the potentially important role of this variable.

Variations in ADC are consistent with the explosiveness of the COVID-19 outbreak in Europe and north-eastern America, where ADC is low, whereas regions with higher ADC have experienced a much slower growth in cases. In particular, Africa and India stand out by high ADC values and low COVID-19 prevalence (Fig. 4-a, Fig. 5). A recent study argued for a reduced transmission rate in Africa potentially linked to the environment, consistent with its higher ADC.^4^ Admittedly, COVID-19 data is quite limited, and very much impacted by policy measures taken to limit disease spread. In addition, testing has been inconsistent across the world; in many countries, reported cases largely refer to individuals showing visible symptoms of the disease, leaving out many asymptomatic cases. Similarly, influenza data is not free from biases (see Methods). This should make us careful in drawing final conclusions. Still, average influenza and COVID-19 prevalence show a similar and consistent relationship to ADC (Fig. 3). Since the high seasonality of influenza is coherent with that of ADC (Figs. 3-c), this suggests that COVID-19 may also follow ADC seasonality, with potential implications for the current disease hotspots of Europe and north-eastern America, where ADC will increase as summer approaches (Fig. 5). In regions of Asia outside India, where the seasonality of ADC is very limited, environmental determinants will probably not play much of a role in shaping COVID-19 dynamics in the months to come. However, the situation may be more worrying in India and Western Africa, two regions where the summer monsoonal systems will bring low ADC conditions offering favourable conditions for the spread of the disease if effective preventive measures are not taken.

**Figure 5.**
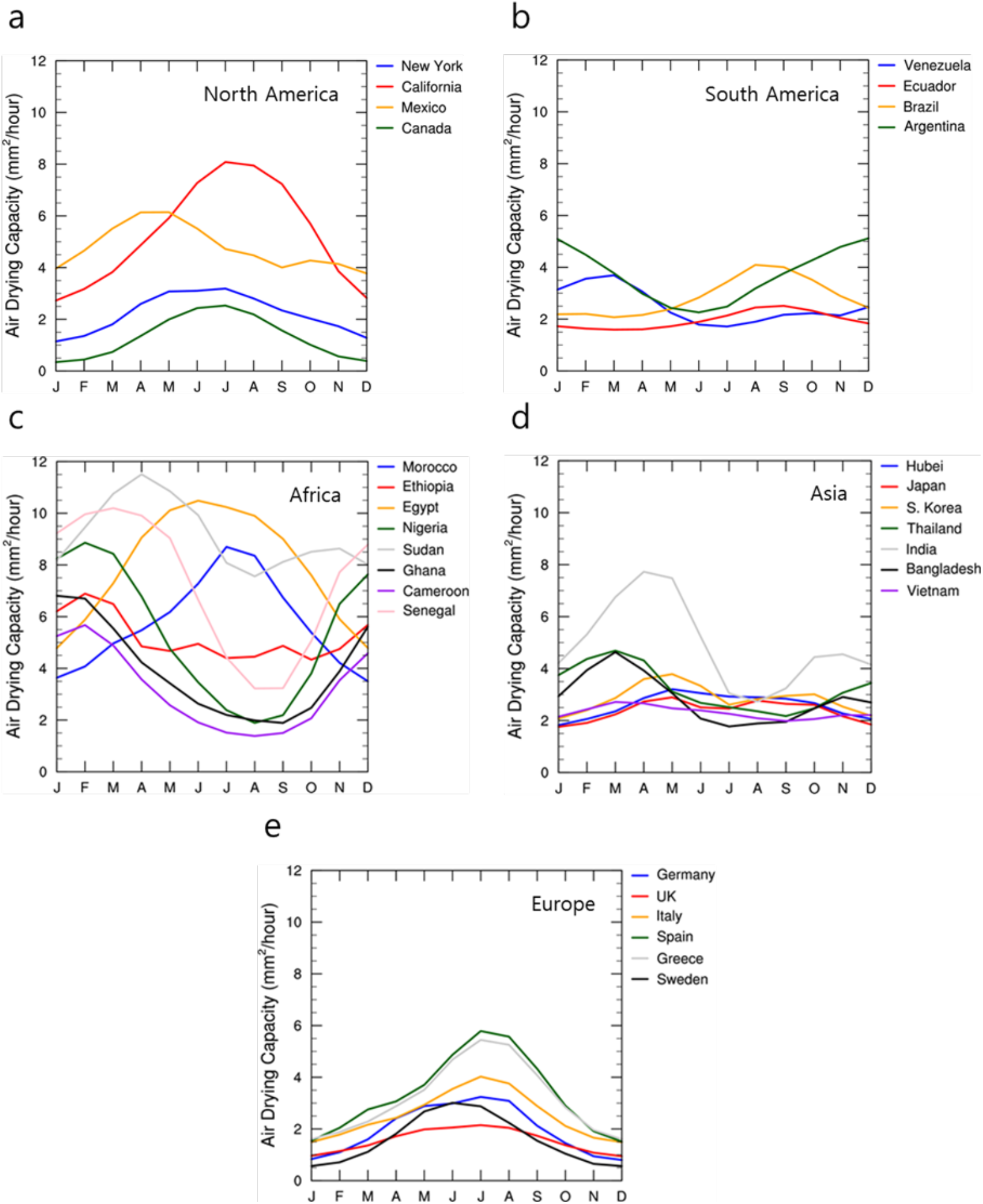
Seasonal variations of ADC. (a-e) Monthly seasonal cycle of ADC for (a) NorthAmerica, (b) South America, (c) Africa, (d) Asia, and (e) Europe. Data from 6-hourly ERA-Interim reanalysis.

Nevertheless, our results present some important caveats. First, indoor heating and cooling will substantially move ADC away from its outdoor value, which we considered in our analysis. Transmission can occur indoors where temperature can be very different from outdoor conditions. Typically, in mid-latitudes, wintertime ADC is much higher inside than outside, and vice-versa during summer. Still, in regions where air conditioning and heating are available, conditions indoors should tend to exhibit much less seasonality than outdoors. In addition, the evident seasonality of influenza makes a strong case for the role of outdoor conditions, given that people spend much of their time indoors year-round^36^. The seasonality of VRDs may therefore primarily reflect outdoor ADC.

Second, biological determinants of virus survival may be strongly correlated to ADC, meaning that part of the ADC-VRD prevalence relationship may be explained by the effect of environmental conditions on the virus itself, and not on the transmission pathway. In particular, temperature is thought to affect the survival of influenza viruses^5^, though we fail to find a coherent signal in global data (Fig. 3-b). Similarly, in the case of influenza and, possibly, COVID-19, UV radiation is believed to be severely detrimental to viruses.^9^ Low ADC is unmistakably associated with low incoming UV, but at higher levels the relationship becomes less clear (Fig. S7). Therefore, ADC and UV radiation may well interact and strengthen their respective effects.

As the COVID-19 pandemic progresses, better data will become available, and it will become possible to test for the robustness of the relationship between its prevalence and ADC values. So far, evidence points to an influenza-like behaviour, with a pronounced seasonality and mid-latitude countries most at risk from late fall to early spring. For the latter, environmental conditions will therefore probably be conducive to a second wave in late 2020, while in Western Africa and India, summer 2020 may bring about favourable conditions for efficient spread of the disease. However, as stressed earlier, conducive environmental conditions are not sufficient to cause VRD spread, and significant outbreaks triggered by social behaviour can occur even under relatively unfavourable environmental conditions.

## Data Availability

6-hourly temperature, dew point temperature and surface pressure data at 0.75 degree spatial resolution between 1979 and 2018 were obtained from the ERA-Interim reanalysis (available at http://apps.ecmwf.int/datasets/). Since ERA-Interim is only available up to 2019, we used 6-hourly ERA5T35 data (0.25 by 0.25 degree horizontal resolution) for the recent February-April 2020 period.
Daily COVID-19 epidemiological data compiled by the Johns Hopkins University Center for Systems Science and Engineering is available at country-scale since 22 January 2020 at https://data.humdata.org/. The most up-to-date COVID data for each US state at a daily temporal resolution is taken from the COVID Tracking Project (https://covidtracking.com/data/). Population data for world countries and US states was downloaded from https://www.worldometers.info/world-population and https://www.wikipedia.org, respectively.
Weekly laboratory confirmed influenza cases by country for the period October 15th, 1995 to August 31st, 2019 are retrieved from the World Health Organization's FluNet database, accessible at https://www.who.int/influenza/gisrs_laboratory/flunet/en/. The dataset consists in weekly totals of identified influenza A and B cases, along with the number of subjects tested.

https://data.humdata.org/

https://covidtracking.com/data/

https://www.who.int/influenza/gisrs_laboratory/flunet/en/

## Author Contributions

E. A. B. E. devised and supervised the study. Y. C. and A. T. carried out analyses. All authors contributed to the manuscript.

## Correspondence

Correspondence and requests for materials should be addressed to.

## References

1. Tamerius JD, Shaman J, Alonso WJ, et al. Environmental Predictors of Seasonal Influenza Epidemics across Temperate and Tropical Climates. PLoS Pathog 2013; 9: e1003194.

2. Araujo MB, Naimi B. Spread of SARS-CoV-2 Coronavirus likely to be constrained by climate. medRxiv 2020; published online Apr 7. DOI:10.1101/2020.03.12.20034728 (preprint).

3. Bukhari Q, Jameel Y. Will Coronavirus Pandemic Diminish by Summer? SSRN 2020; published online Mar 17. DOI:10.2139/ssrn.3556998 (preprint).

4. Cabore JW, Karamagi H, Kipruto H, et al. The potential effects of widespread community transmission of SARS-CoV-2 infection in the WHO African Region: a predictive model. BMJ global health 2020; accepted (https://gh.bmj.com/pages/wp-content/uploads/sites/58/2020/05/BMJGH-The_potential_effects_of_widespread_community_transmission_of_SARS-CoV-2_infection_in_the_WHO_African_Region_a_predictive_model-Copy.pdf).

5. Polozov IV., Bezrukov L, Gawrisch K, Zimmerberg J. Progressive ordering with decreasing temperature of the phospholipids of influenza virus. Nat Chem Biol 2008; 4: 248–255.

6. Shaman J, Kohn M. Absolute humidity modulates influenza survival, transmission, and seasonality. Proc Natl Acad Sci USA 2009; 106: 3243–3248.

7. Wang J, Tang K, Feng K, Lv W. High Temperature and High Humidity Reduce the Transmission of COVID-19. SSRN 2020; published online Mar 10. DOI:10.2139/ssrn.3551767 (preprint).

8. Chin AWH, Chu JTS, Perera MRA, et al. Stability of SARS-CoV-2 in different environmental conditions. medRxiv 2020; published online Mar 27. DOI:10.1101/2020.03.15.20036673 (preprint).

9. Sagripanti JL, Lytle CD. Inactivation of influenza virus by solar radiation. Photochem Photobiol 2007; 83: 1278–1282.

10. Homeland Security Science and Technology. Response to SARS-CoV-2 / COVID-19.Apr 20, 2020.https://www.dhs.gov/sites/default/files/publications/panthr_covid-19_fact_sheet_v13_27apr-final_0.pdf (accessed May 17, 2020).

11. Lowen AC, Mubareka S, Steel J, Palese P. Influenza virus transmission is dependent on relative humidity and temperature. PLoS Pathog 2007; 3: 1470–1476.

12. Xie X, Li Y, Chwang ATY, Ho PL, Seto WH. How far droplets can move in indoor environments – revisiting the Wells evaporation-falling curve. Indoor Air 2007; 17: 211–25.

13. Ishmatov A. Influence of weather and seasonal variations in temperature and humidity on supersaturation and enhanced deposition of submicron aerosols in the human respiratory tract. Atmos Environ 2020; 17: 211–25.

14. Deyle ER, Maher MC, Hernandez RD, Basu S, Sugihara G. Global environmental drivers of influenza. Proc Natl Acad Sci USA 2016; 113: 13081–13086.

15. Ficetola GF, Rubolini D. Climate affects global patterns of COVID-19 early outbreak dynamics. medRxiv 2020; published online Apr 20. DOI:10.1101/2020.03.23.20040501 (preprint).

16. Sajadi MM, Habibzadeh P, Vintzileos A, Shokouhi S, Miralles-Wilhelm F, Amoroso A. Temperature, humidity, and latitude analysis to predict potential spread and seasonality for COVID-19. SSRN 2020; published online Apr 6. DOI:10.2139/ssrn.3550308 (preprint).

17. Luo W, Majumder MS, Liu D, et al. The role of absolute humidity on transmission rates of the COVID-19 outbreak. medRxiv 2020; published online Feb 17. DOI:10.1101/2020.02.12.20022467 (preprint).

18. Poirier C, Luo W, Majumder M, et al. The Role of Environmental Factors on Transmission Rates of the COVID-19 Outbreak: An Initial Assessment in Two Spatial Scales. SSRN 2020; published online Apr 16. DOI:10.2139/ssrn.3552677 (preprint).

19. Atkinson MP, Wein LM. Quantifying the routes of transmission for pandemic influenza. Bull Math Biol 2008; 70: 820–867.

20. Stilianakis NI, Drossinos Y. Dynamics of infectious disease transmission by inhalable respiratory droplets. J R Soc Interface 2010; 1355–1366.

21. Cowling BJ, Ip DKM, Fang VJ, et al. Aerosol transmission is an important mode of influenza A virus spread. Nat Commun 2013; 4: 1935.

22. Smieszek T, Lazzari G, Salathé M.. Assessing the Dynamics and Control of Droplet-and Aerosol-Transmitted Influenza Using an Indoor Positioning System. Sci Rep 2019; 9: 2185.

23. Asadi S, Wexler AS, Cappa CD, Barreda S, Bouvier NM, Ristenpart WD. Aerosol emission and superemission during human speech increase with voice loudness. Sci Rep 2019; 9: 2348.

24. Han ZY, Weng WG, Huang QY. Characterizations of particle size distribution of the droplets exhaled by sneeze. J R Soc Interface 2013; 10: 20130560.

25. Wells WF. On air-borne infection: Study II. Droplets and droplet nuclei. Am J Epidemiol 1934; 20: 611–618.

26. Bourouiba L. Turbulent Gas Clouds and Respiratory Pathogen Emissions: Potential Implications for Reducing Transmission of COVID-19. JAMA – J Am Med Assoc 2020; 323: 1837–1838.

27. WHO. Infection prevention and control of epidemic-and pandemic-prone acute respiratory infections in health care. Apr, 2014. https://www.who.int/csr/bioriskreduction/infection_control/publication/en/ (accessed May 17, 2020).

28. Siegel JD, Rhinehart E, Jackson M, Chiarello L. Guideline for isolation precautions: Preventing transmission of infectious agents in healthcare settings. 2007. https://www.cdc.gov/infectioncontrol/guidelines/isolation/index.html (accessed May 17. 2020)

29. Bourouiba L, Dehandschoewercker E, Bush JWM. Violent respiratory events: On coughing and sneezing. J Fluid Mech 2014; 745: 537–563.

30. WHO. Report of the WHO–China Joint Mission on Coronavirus Disease2019 (COVID-19), 16–24 Feb, 2020. https://www.who.int/docs/default-source/coronaviruse/who-china-joint-mission-on-covid-19-final-report.pdf (accessed May 17. 2020).

31. Liu Y, Ning Z, Chen Y, et al. Aerodynamic analysis of SARS-CoV-2 in two Wuhan hospitals. Nature 2020; published online Apr 27. DOI: 10.1038/s41586-020-2271-3 (preprint).

32. Maxwell JC. The Scientific Papers of James Clerk Maxwell Vol. 1 (Dover, New York, 2003)

33. Rogers RR, Yau MK. A Short Course in Cloud Physics (Pergamon, 1989).

34. Dee DP, Uppala SM, Simmons AJ, et al. The ERA-Interim reanalysis: Configuration and performance of the data assimilation system. Q J R Meteorol Soc 2011; 137: 553–597.

35. Hersbach H, De Rosnay P, Bell B, et al. Operational global reanalysis: progress, future directions and synergies with NWP including updates on the ERA5 production status. ERA Rep Ser 2018. https://www.ecmwf.int/en/elibrary/18765-operational-global-reanalysis-progress-future-directions-and-synergies-nwp (accessed May 17. 2020).

36. Tamerius J, Nelson MI, Zhou SZ, Viboud C, Miller MA, Alonso WJ. Global influenza seasonality: Reconciling patterns across temperate and tropical regions. Environ Health Perspect 2011; 119: 439–445.

37. Effros, R. M. et al. Dilution of respiratory solutes in exhaled condensates. Am. J. Respir. Crit. Care Med. 2002 165: 663–669.

